# Intra-arterial recombinant human TNK tissue-type plasminogen activator (rhTNK-tPA) thrombolysis for acute medium vessel occlusion (MeVO-TNK): Study rationale and design

**DOI:** 10.64898/2026.06.16.26355640

**Authors:** Gang Deng, Chen-Chen Liu, Yi-Hui Wang, Dong-Hui Ao, Hao Huang, Yi Xie, Yi Zhang, Qian-Qian Kong, Ling-Ning Lan, Pei-Xin Li, Ting-Ting Qin, Guo Li, Sha-Bei Xu, Xiang Luo

## Abstract

**Background:** The optimal management of acute ischemic stroke caused by medium vessel occlusion (MeVO) remains uncertain. Recent randomized trials have failed to demonstrate a clear benefit of endovascular therapy in this population, whereas intra–arterial thrombolysis (IAT) has emerged as a biologically plausible alternative. However, prospective evidence supporting IAT in MeVO is lacking, and the optimal dosing strategy for stand–alone IAT remains undefined.

**Aim:** To preliminarily evaluate the efficacy and safety of intra–arterial tenecteplase (IA–TNK) plus standard medical therapy (SMT) compared with SMT alone in patients with acute MeVO stroke, and to explore a stepwise IA–TNK dosing strategy.

**Design:** The MeVO–TNK trial is a multicenter, prospective, randomized, open–label, blinded–endpoint (PROBE), exploratory phase II study. A total of 60 participants with imaging–confirmed MeVO will be randomized 1:1 to receive either IA–TNK plus SMT or SMT alone. Participants presenting beyond 6 hours from symptom onset must demonstrate salvageable penumbral tissue on advanced imaging. Those assigned to the intervention group will receive up to two intra–arterial boluses of tenecteplase (0.0625 mg/kg per bolus), with the second bolus administered based on angiographic assessment of reperfusion and safety.

**Outcomes:** The primary efficacy outcome is final infarct volume measured at 72 ± 24 hours after randomization. Secondary efficacy outcomes include the proportions of patients achieving modified Rankin Scale (mRS) scores of 0–1, 0–2 and 0–3 at 90 days, a shift analysis of the mRS distribution at 90 days, early neurological deterioration, and National Institutes of Health Stroke Scale score at 7 days or discharge. The primary safety outcome is symptomatic intracranial hemorrhage within 24 hours.

**Conclusions:** This trial will provide preliminary evidence on the biological efficacy, reperfusion potential and safety of stand–alone IA–TNK for acute MeVO stroke, helping to address an important evidence gap and inform the design of future confirmatory studies.

**Trial Registration:** Chinese Clinical Trial Registry, ChiCTR2400091249.

**WHAT IS ALREADY KNOWN ON THIS TOPIC:** The optimal reperfusion strategy for medium vessel occlusion (MeVO) remains uncertain, and no randomized trial has specifically evaluated intra-arterial tenecteplase (IA-TNK) as a stand-alone treatment for acute MeVO stroke.

**WHAT THIS STUDY ADDS:** The MeVO-TNK trial is a multicenter randomized study that will assess the efficacy and safety of IA-TNK plus standard medical therapy versus standard medical therapy alone and explore a stepwise IA-TNK dosing strategy.

**HOW THIS STUDY MIGHT AFFECT RESEARCH, PRACTICE OR POLICY:** The findings will provide preliminary evidence to guide future definitive trials and may support the development of alternative reperfusion strategies for patients with MeVO.

## Introduction and rationale

Intracranial medium vessel occlusion (MeVO) accounts for approximately one-quarter to two-fifths of acute ischemic strokes (AIS),^1,2^ yet the optimal treatment strategy remains uncertain. Approximately one-third of patients fail to achieve functional independence and nearly 10% die within 90 days despite contemporary medical management, including intravenous thrombolysis (IVT) in eligible patients.^3^ Furthermore, although IVT may achieve recanalization in a subset of patients, recanalization rates remain modest, leaving a substantial proportion of patients with persistent vessel occlusion.^3,4^ Recent randomized controlled trials evaluating endovascular therapy (EVT) for MeVO have yielded mixed results, with several studies failing to demonstrate a clear clinical benefit over best medical management.^5,6^ Furthermore, these studies underscored the challenges of achieving a favorable benefit–risk balance in this population, particularly given the potential for procedure-related complications and intracranial hemorrhage associated with endovascular intervention. These findings challenge the routine application of EVT in MeVO and underscore the need for alternative reperfusion strategies that can balance efficacy with safety.

Compared with EVT, intra-arterial thrombolysis (IAT) offers several potential advantages for the treatment of MeVO. IAT avoids mechanical manipulation of fragile distal cerebral vessels, thereby potentially reducing endothelial injury and procedure-related complications. In addition, direct delivery of a thrombolytic agent to the occlusion site may facilitate effective clot dissolution while maintaining procedural simplicity and improving access to distal and tortuous vascular territories.^7^ Supporting this concept, a post hoc subgroup analysis of the PROACT-II trial demonstrated improved reperfusion and functional outcomes with intra-arterial prourokinase in patients with isolated M2 occlusions.^8^ Furthermore, a retrospective study evaluating rescue IAT with urokinase following failed or incomplete mechanical thrombectomy reported improved reperfusion and functional outcomes without an increase in symptomatic intracranial hemorrhage (sICH) or mortality.^9^ Collectively, these findings suggest that IAT may represent a promising therapeutic option for selected patients with MeVO.

Tenecteplase (TNK) is a genetically modified variant of alteplase characterized by greater fibrin specificity, increased resistance to plasminogen activator inhibitor-1, and a longer plasma half-life.^10^ These pharmacological properties make TNK an attractive candidate for intra-arterial administration, potentially enabling more efficient thrombus dissolution while limiting systemic fibrinolytic effects. Reflecting these advantages, TNK has been incorporated into contemporary stroke guidelines in several countries as an alternative to alteplase for intravenous thrombolysis.^11,12^ However, despite its established role in IVT and its favorable pharmacological profile, no prospective clinical study has specifically evaluated intra-arterial TNK (IA-TNK) as a primary reperfusion strategy for MeVO. Notably, a recent international survey of stroke interventionists demonstrated substantial interest in this approach, with 76.5% of respondents indicating willingness to use IA-TNK in the treatment of MeVO.^13^

Therefore, the MeVO-TNK trial was designed as a multicenter, prospective, randomized, open-label, blinded-endpoint pilot study to preliminarily evaluate the efficacy and safety of IA-TNK compared with standard medical treatment (SMT) in patients with acute ischemic stroke caused by MeVO.

## Methods

### Study design

The MeVO-TNK trial is an investigator-initiated, multicenter, prospective, randomized, open-label, blinded-endpoint clinical trial with a PROBE (Prospective Randomized Open, Blinded Endpoint) design. The study was registered in the Chinese Clinical Trial Registry (ChiCTR***) before participant enrollment and is being conducted at 14 comprehensive stroke centers in China.

The trial was designed and conducted in accordance with the Declaration of Helsinki, the International Council for Harmonisation Good Clinical Practice (ICH-GCP) guidelines, and applicable national regulations. The protocol was approved by the Ethics Committee of *** (No. ***) and by the institutional review board of each participating center.

Potentially eligible participants undergo diagnostic cerebral angiography. Randomization is performed only after angiographic confirmation of the target MeVO and verification of all eligibility criteria. Participants are randomized in a 1:1 ratio to receive either intra-arterial tenecteplase plus standard medical therapy or standard medical therapy alone.

Treatment allocation is open label; however, all efficacy and safety outcomes are assessed by independent investigators and core laboratories blinded to treatment assignment. Neuroimaging outcomes are centrally adjudicated by an independent Imaging Core Laboratory, and functional outcomes are assessed by trained evaluators who remain unaware of treatment allocation throughout follow-up.

The original protocol (Version 1.0, August 18, 2024) restricted enrollment to participants presenting between 4.5 and 24 hours after symptom onset or last known well and excluded those who had received intravenous thrombolysis (IVT). Following review of emerging evidence and contemporary stroke treatment guidelines, Protocol Version 2.0 was approved on February 14, 2025, expanding eligibility to participants presenting within 24 hours and permitting prior IVT according to guideline recommendations. These amendments were approved by the ethics committees of all participating centers and implemented after updating the trial registry.

The overall trial flowchart is shown in **Figure 1**, and the schedule of enrollment, interventions, and assessments is provided in Supplemental **Table 1**.

**Figure 1.**
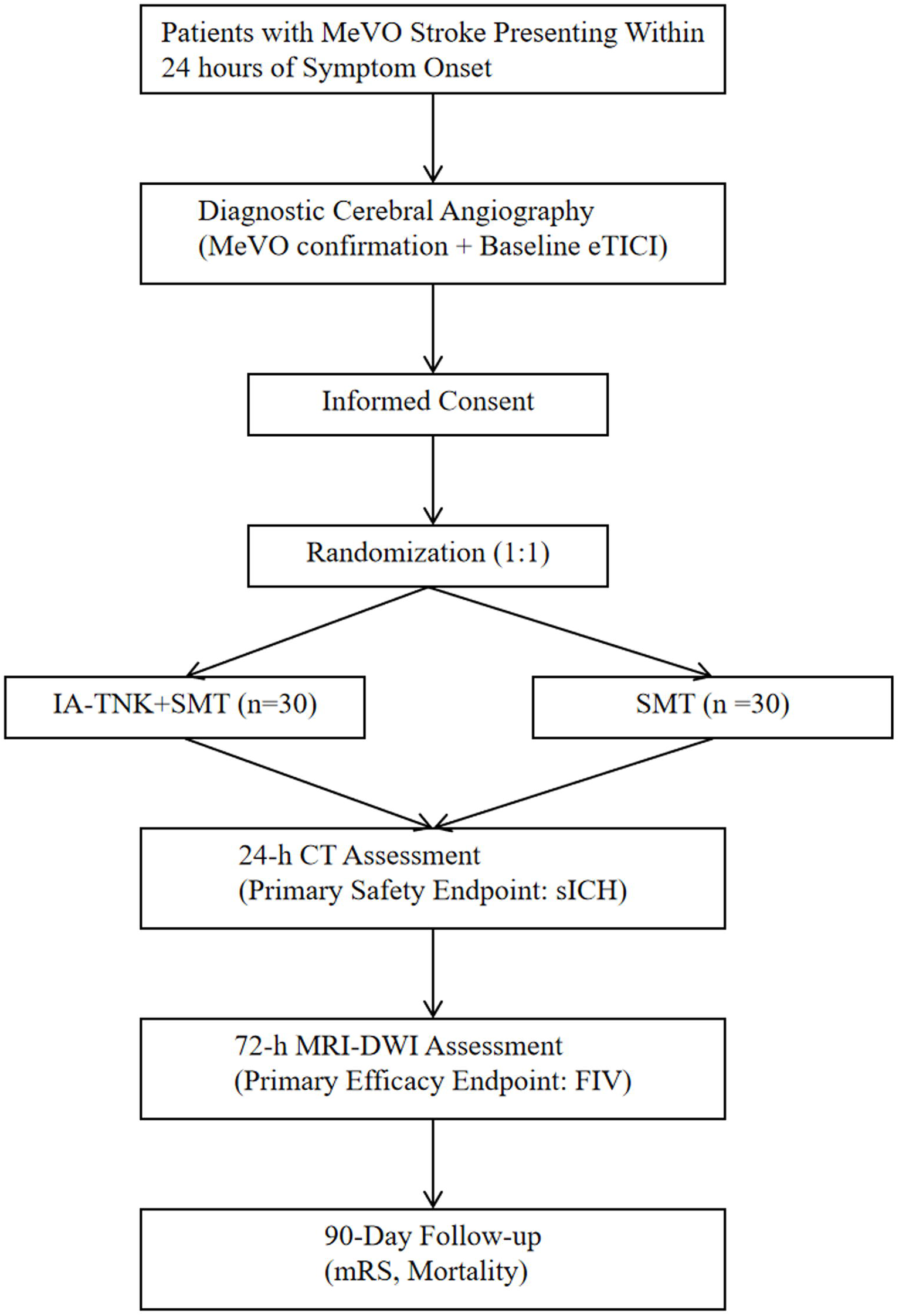
Flow chart of the study. MeVO, medium vessel occlusion; eTICI, extended Thrombolysis In Cerebral Infarction; IA-TNK, intra-arterial tenecteplase; SMT, standard medical treatment; sICH, symptomatic intracranial hemorrhage; MRI-DWI, magnetic resonance imaging with diffusion-weighted imaging; FIV, final infarct volume; mRS, modified Rankin score.

### Eligibility criteria

Eligible participants were adults with imaging-confirmed acute isolated MeVO who could be randomized within 24 hours of symptom onset or last known well. For participants presenting beyond 6 hours, evidence of salvageable brain tissue on advanced imaging was required. Key exclusion criteria included pre-stroke disability, intracranial hemorrhage on baseline imaging, planned mechanical thrombectomy, and contraindications to thrombolytic therapy. Detailed inclusion and exclusion criteria are provided in **Figure 2**.

**Figure 2.**
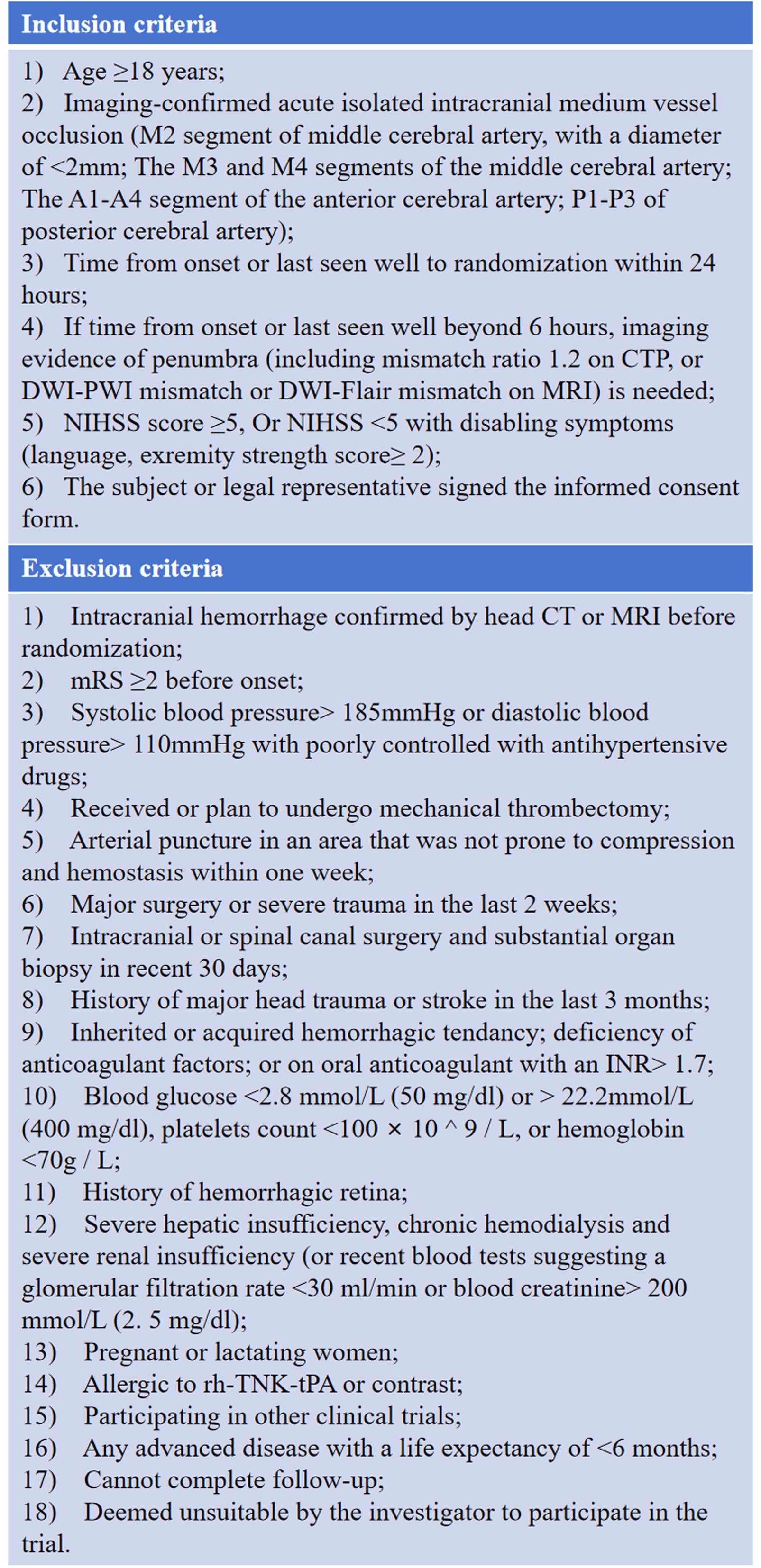
Inclusion and exclusion criteria. CTP, computed tomography perfusion; DWI, diffusion-weighted imaging; PWI, perfusion-weighted imaging; Flair, fluid-attenuated inversion recovery; MRI, magnetic resonance imaging; NIHSS, National Institutes of Health Stroke Scale; mRS, modified Rankin score; rh-TNK-tPA, recombinant human TNK tissue-type plasminogen activator.

### Randomization

A centralized, web-based Electronic Data Capture (EDC) and Interactive Web Response System (IWRS) (https://edc-cloud.medsci.cn/), provided by Shanghai MedSci Co., Ltd, will manage study data collection and treatment allocation.

Following diagnostic cerebral angiography and confirmation of all eligibility criteria, participants will be randomized in a 1:1 ratio to either the IA-TNK group or the SMT group.

Randomization will be performed centrally through the IWRS using a permuted block randomization scheme with randomly varying block sizes of 2 and 4 to maintain allocation concealment. Randomization will not be stratified by study center because of the relatively small number of participants expected at each site.

The randomization sequence will be generated by an independent statistician using SAS software (version 9.4) and implemented through the IWRS. Treatment allocation will be revealed to the treating investigators after randomization. Although treatment assignment is open label, all efficacy and safety endpoint assessments will be performed by independent evaluators blinded to treatment allocation.

### Treatments

All potentially eligible patients underwent diagnostic cerebral angiography before randomization. IVT before randomization was permitted when administered according to current guideline recommendations.

Patients randomized to the IA-TNK group received IA-TNK in addition to SMT. A microcatheter was advanced to the proximal end of the target occlusion through a guiding or intermediate catheter. TNK was prepared as a 1.33 mg/mL solution and administered intra-arterially at an initial dose of 0.0625 mg/kg (maximum dose, 6.25 mg) at a constant infusion rate of 1 mL/min.

Five minutes after completion of the infusion, repeat angiography was performed and the expanded Thrombolysis in Cerebral Infarction (eTICI) score was assessed. If angiographic reperfusion reached eTICI 2b50 or higher, or if contrast extravasation suggestive of hemorrhage was observed on control angiography, no further TNK was administered. Otherwise, a second identical dose of intra-arterial TNK was administered at the same infusion rate. A final angiographic assessment was performed 5 minutes after completion of the second infusion.

Participants randomized to the control group received SMT alone. SMT was provided according to contemporary guideline-based management of AIS and local institutional practice. No additional intra-arterial thrombolytic agents were permitted after randomization. Mechanical thrombectomy or any other endovascular reperfusion treatment was not permitted after randomization in either treatment group.

### Imaging Protocol and Infarct Volume Assessment

All imaging assessments are performed according to a standardized protocol. Baseline infarct core volume is estimated using imaging modality–specific criteria. On CT perfusion (CTP), ischemic core is defined as regions with relative cerebral blood flow (rCBF) <30% compared with the contralateral hemisphere. On MRI, infarct core is defined using diffusion-weighted imaging (DWI) with apparent diffusion coefficient (ADC)-based thresholds consistent with established literature.

In participants evaluated only with non-contrast CT without advanced imaging, baseline infarct core volume is considered missing for primary adjusted analyses. Missing baseline infarct core volume data will be handled using multiple imputation in the primary intention-to-treat analysis. Sensitivity analyses using complete-case and per-protocol populations will be performed to assess the robustness of the findings.

The primary endpoint, final infarct volume (FIV), is assessed at 72 ± 24 hours after randomization, preferentially using MRI-DWI. If MRI is unavailable, CT acquired within the same time window is used. All imaging data are centrally adjudicated by a blinded Imaging Core Laboratory and processed using a standardized workflow on the UII AI Scientific Research Platform (https://www.uiiai.com/en/uai/scientificresearch). All analysts remain blinded to treatment allocation and clinical outcomes.

### Efficacy endpoints

The primary efficacy endpoint is the FIV at 72 ± 24 hours after randomization. Secondary efficacy endpoints include

1. Proportion of participants with no disability (modified Rankin Scale [mRS] score 0–1) at 90 (±7) days;
2. Proportion of participants with functional independence (mRS score 0–2) at 90 (±7) days;
3. Proportion of participants with independent ambulation (mRS score 0–3) at 90 (±7) days;
4. Distribution of mRS scores at 90 (±7) days (shift analysis);
5. Early neurological deterioration, defined as an increase in NIHSS score of ≥2 points from baseline to 72 ± 24 hours;
6. NIHSS score at 7 ± 1 days after randomization or at hospital discharge, whichever occurs first.

### Safety endpoints

The primary safety endpoint is sICH within 24 hours after randomization, defined according to the Heidelberg Bleeding Classification.^14^

Secondary safety endpoints include

1. All-cause mortality within 90 (±7) days after randomization;
2. Any intracranial hemorrhage occurring within 72 hours after randomization, regardless of clinical symptoms.
3. Adverse events (AEs) and serious adverse events (SAEs)

All intracranial hemorrhage events will be adjudicated by an independent Imaging Core Laboratory and the Clinical Events Committee blinded to treatment allocation.

### Data and Safety Monitoring Board

An independent Data and Safety Monitoring Board (DSMB) will be established to oversee the trial. The DSMB will comprise three experts: a neurologist, a neurointerventionalist, and a biostatistician, all independent of the study investigators and the sponsor. Members of the DSMB will have no involvement in patient enrollment, treatment administration, or outcome assessments. The DSMB will meet at regular intervals, at least annually, to review trial conduct, enrollment progress, protocol adherence, and safety data, including the occurrence of serious adverse events. Adverse events will be evaluated using the fifth edition of the Common Terminology Criteria for Adverse Events of the USA.^15^ Based on these reviews, the DSMB may provide recommendations regarding the continuation, modification, or termination of the trial to ensure participant safety.

### Sample size estimate

Given the exploratory nature of this phase II study, the sample size was not based on formal hypothesis testing. Instead, 60 participants (30 per group) were considered adequate to assess feasibility, obtain preliminary estimates of efficacy and safety, and inform the design of a future definitive trial. This sample size is consistent with published recommendations for pilot randomized studies and with the sample sizes commonly observed in contemporary pilot and feasibility trials.^16,17^

### Statistical analysis

All statistical analyses will be conducted using SAS version 9.4 (SAS Institute Inc) and R version 4.3.0 (R Core Team). Continuous variables will be summarized as mean (standard deviation) or median (interquartile range), as appropriate, and categorical variables as counts and percentages. Statistical tests will be two-sided, with P < 0.05 considered statistically significant, and estimates of treatment effect will be reported with 95% confidence intervals (CIs).

The primary efficacy analysis will be performed in the intention-to-treat (ITT) population, defined as all randomized participants regardless of treatment received. Safety analyses will be performed in the safety population, including all participants who received any study treatment.

The primary endpoint will be analyzed using a generalized linear model (GLM) with a Gamma distribution and log link, adjusting for baseline infarct core volume. Treatment effects will be expressed as adjusted mean ratios with 95% CIs.

Missing primary outcome data will not be imputed, and the primary analysis will be conducted using all available FIV data. Because baseline infarct core volume is included as an adjustment variable in the primary model, missing baseline infarct core volume values will be handled using multiple imputation under the assumption that data are missing at random. Prespecified sensitivity analyses will include analysis in the per-protocol population (participants who adhered to the study protocol without major deviations) and repetition of the primary model after excluding participants with missing baseline infarct core volume.

Secondary efficacy outcomes will be analyzed using GLM or appropriate nonparametric methods, adjusting for relevant baseline covariates where applicable. Functional outcomes based on the mRS will be adjusted for age and baseline NIHSS score, whereas analyses of NIHSS score and early neurological deterioration will be adjusted for baseline NIHSS score.

An exploratory subgroup analysis will assess the consistency of treatment effects according to prior IVT status (yes vs no). Potential treatment-by-subgroup interactions may be explored within the primary analysis model. Given the exploratory nature of the study and limited sample size, subgroup findings will be interpreted cautiously.

Exploratory analyses will assess infarct growth, defined as 72-hour FIV minus baseline infarct core volume; angiographic reperfusion efficacy, including change in eTICI score and achievement of eTICI ≥2b50; and the potential influence of prior IVT on efficacy and safety outcomes. Additional exploratory analyses may evaluate the consistency of treatment effects across clinically relevant participant subgroups, including age, baseline NIHSS score, baseline infarct core volume, time from symptom onset to randomization, prior IVT status, and occlusion location. These analyses are intended to generate hypotheses for future studies and will be interpreted cautiously. Exploratory endpoints will be summarized descriptively, and associations may be explored using GLM or other appropriate regression methods adjusted for relevant baseline covariates.

No adjustment for multiplicity will be applied to secondary or exploratory analyses, and these results will be considered hypothesis-generating.

Categorical safety outcomes will be summarized as frequencies and percentages. Relative risks with 95% CIs for sICH and any intracranial hemorrhage will be estimated using modified Poisson regression. Time-to-event outcomes will be analyzed using Kaplan–Meier methods, log-rank tests, and Cox proportional hazards models, as appropriate.

### Ethics and Dissemination

The trial will be conducted in accordance with the Declaration of Helsinki, the International Council for Harmonisation Good Clinical Practice (ICH-GCP) guidelines, and applicable national regulations. Ethical approval was obtained from the Ethics Committee of Tongji Medical College, Huazhong University of Science and Technology (No. 2024-S053) and from the institutional review boards of all participating centers. Written informed consent will be obtained from all participants or their legally authorized representatives before enrollment.

Participant confidentiality will be protected through the use of unique study identification numbers and secure electronic data storage with restricted access. Insurance coverage for trial-related harm will be provided in accordance with applicable regulations. The study findings will be disseminated through presentations at scientific meetings and publication in peer-reviewed journals regardless of the study outcomes.

### Trial status

This study was registered on October 23, 2024, and recruitment began on November 21, 2024. The enrollment was finished on July, 2025.

## Discussion

MeVO accounts for a substantial proportion of acute ischemic strokes and is associated with suboptimal clinical outcomes under current standard therapy.^1,2^ Although neurological deficits are generally less severe than those observed in large vessel occlusion, a considerable proportion of patients fail to achieve functional independence.^3^ Recent randomized controlled trials evaluating EVT for MeVO have yielded conflicting results. While the DISTAL, ESCAPE-MeVO, and DISCOUNT trials did not demonstrate a clear clinical benefit and raised concerns regarding procedural risks,^5,6^ the recently published ORIENTAL-MeVO trial reported improved functional outcomes with EVT in selected patients.^18^ Collectively, these findings suggest that the optimal reperfusion strategy for MeVO remains uncertain and may depend on patient selection, occlusion characteristics, and procedural factors. In this context, IAT represents a biologically plausible and potentially less invasive alternative, particularly for distal or tortuous occlusions where mechanical approaches may be technically challenging. However, prospective evidence supporting IAT in MeVO remains scarce.

Tenecteplase (TNK), a genetically modified tissue plasminogen activator, has several pharmacological advantages over alteplase, including higher fibrin specificity, increased resistance to plasminogen activator inhibitor-1, and a longer plasma half-life. These properties make TNK an attractive candidate for intra-arterial administration in MeVO. However, there is currently a lack of prospective clinical evidence evaluating stand-alone intra-arterial TNK in this population, particularly with respect to optimal dosing, safety, and efficacy. Several recent studies have provided important insights into the adjunctive intra-arterial use of tenecteplase in AIS due to LVO. The ANGEL-TNK trial demonstrated that adjunctive IA-TNK administered after successful thrombectomy improved 90-day functional outcomes without increasing the risk of sICH.^19^ In contrast, the POST-TNK and ATTENTION-IA trials reported neutral efficacy results while confirming the safety of a lower IA-TNK dose (0.0625 mg/kg) in patients with successful reperfusion.^20,21^ Furthermore, the DATE study identified 0.0625 mg/kg as the dose with the most favorable balance between efficacy and safety signals for post-EVT microcirculatory augmentation.^22^ Collectively, these studies support the biological activity and safety of IA-TNK, while highlighting the lack of evidence regarding its use as a stand-alone reperfusion therapy in patients with MeVO.

The MeVO-TNK trial was designed to address this gap. This prospective, multicenter, randomized study will enroll 60 participants to compare intra-arterial TNK with standard medical therapy. The trial employs a predefined stepwise dosing strategy, administering an initial bolus of 0.0625 mg/kg with a conditional second bolus based on angiographic results, to balance safety and therapeutic efficacy. For patients presenting beyond six hours from symptom onset, advanced imaging is required to confirm salvageable penumbral tissue, thereby enriching the study population with patients most likely to benefit from reperfusion. Importantly, the study uses final infarct volume at 72 ± 24 hours as the primary endpoint, which is sensitive to biological treatment effects and suitable for an exploratory phase II study with a limited sample size.

During the study, the protocol was amended to allow inclusion of patients who had received intravenous thrombolysis and to expand the enrollment window to 24 hours from symptom onset, improving recruitment feasibility and generalizability while maintaining scientific integrity. This flexibility ensures that the trial reflects contemporary clinical practice without compromising methodological rigor.

This exploratory trial is expected to provide important insights into the safety and biological efficacy of stand-alone intra-arterial TNK for MeVO. A positive result would support the use of TNK as a novel treatment option in this under-studied population, whereas a negative result would provide critical evidence to guide future investigations toward alternative pharmacological strategies or refined patient selection. Data collected on reperfusion efficacy, treatment timelines, and imaging biomarkers will also contribute to a better understanding of MeVO pathophysiology and inform the design of subsequent trials.

The strengths of this study include its prospective, randomized, multicenter design; imaging-based patient selection; blinded assessment of outcomes via an independent core laboratory; and the use of a mechanistically relevant imaging endpoint.

Several limitations should be acknowledged. First, as an exploratory phase II study, the sample size is not powered to detect modest treatment effects or to support definitive subgroup analyses. Second, the primary endpoint is FIV, an imaging-based surrogate outcome rather than a direct measure of long-term functional recovery. Although infarct volume is biologically relevant and may increase sensitivity for detecting treatment effects in a pilot study, its relationship with clinical outcomes is not absolute. Third, although imaging assessments will be standardized and centrally adjudicated, the use of both MRI-DWI and CT for infarct volume assessment may introduce some degree of measurement heterogeneity. Fourth, treatment allocation is open label, and the inclusion of participants who receive IVT may introduce additional variability in treatment response despite randomization. Finally, the stringent imaging selection criteria and conduct of the study at experienced stroke centers may limit the generalizability of the findings to broader clinical practice.

Overall, the MeVO-TNK trial is designed to provide initial high-quality evidence regarding intra-arterial TNK for acute MeVO, offering valuable guidance for future confirmatory studies and optimized reperfusion strategies.

## Data Availability

All data produced in the present study are available upon reasonable request to the authors

## Acknowledgements

We are grateful to all the participants in the MeVO-TNK program for their contributions to this study.

## Contributors

GD, SBX and XL designed the study; GD, SBX, CCL, YHW, DHA and HH drafted the manuscript; YX, YZ, QQK, LNL, PXL, TTQ, GL provided critical comments/revisions of the manuscript. XL is the guarantor.

## Funding

The trial receives support from the National Natural Science Foundation of China (Grant No. 82171385), the High-Quality Clinical Research Fund of Tongji Hospital (Grant No. 2024TJCR013), and the Hubei Provincial Natural Science Foundation of China (Grant No. 2025AFB718). The funder had no role in study design, data collection, analysis, interpretation, or manuscript writing.

## Competing interests

None declared.

## Patient consent for publication

Not applicable.

## Ethics approval

This study involves human participants and was approved by the ethics committee of Tongji Medical College, Huazhong University of Science and Technology (IRB approval number: 2024-S053). Participants provided informed consent before taking part in the study.

## Notes

### Competing Interest Statement

The authors have declared no competing interest.

### Clinical Trial

Chinese Clinical Trial Registry, ChiCTR2400091249.

### Author Declarations

This study involves human participants and was approved by the ethics committee of Tongji Medical College, Huazhong University of Science and Technology (IRB approval number: 2024-S053).

