## Supplemental table 1 for "Intra-arterial recombinant human TNK tissue-type plasminogen activator (rhTNK-tPA) thrombolysis for acute medium vessel occlusion (MeVO-TNK): Study rationale and design"

**Supplemental table 1** Schedule of enrollment, interventions, and assessments

| **Assessments** | **Visit (Time in hours / days)** | | | | | |
| --- | --- | --- | --- | --- | --- | --- |
|  | **Screening/**  **Randomization** | **Treatment** | **Follow-up** | | | |
|  | **V0** | **V1** | **V2 (24h)** | **V3 (72±24h)** | **V4 (7±1 d or discharge)** | **V5 (90 ±7 d)** |
| Informed consent | X |  |  |  |  |  |
| Eligibility Assessment | X |  |  |  |  |  |
| Demographics | X |  |  |  |  |  |
| Medical History | X |  |  |  |  |  |
| Physical Examination | X |  | X | X | X | X^*^ |
| Vital Signs (BP/HR) | X | X | X | X |  |  |
| Laboratory Tests | X |  | X |  |  |  |
| Pregnancy Test (if applicable) | X |  |  |  |  |  |
| 12-lead ECG | X |  |  |  |  |  |
| Pre-stroke mRS | X |  |  |  |  |  |
| NIHSS | X |  | X | X | X |  |
| CT/MRI Brain Imaging | X |  | X (CT) | X (DWI/CT) |  |  |
| CTA/MRA | X |  |  |  |  |  |
| DSA | X |  |  |  |  |  |
| Baseline eTICI Score | X |  |  |  |  |  |
| IA TNK Administration |  | X |  |  |  |  |
| Post-treatment eTICI Score |  | X |  |  |  |  |
| Concomitant Medications | X | X | X | X | X | X |
| Adverse Events |  | X | X | X | X | X |
| Serious Adverse Events |  | X | X | X | X | X |
| Final Infarct Volume Assessment |  |  |  | X |  |  |
| mRS Assessment |  |  |  |  | X | X |
| Mortality Assessment |  |  |  |  |  | X |

^*^Physical examination at Day 90 will be performed if the participant returns for an on-site visit.

BP, blood pressure; HR, heart rate; ECG, electrocardiogram; mRS, modified Rankin scale; NIHSS, National Institutes of Health Stroke Scale; CT, computed tomography; MRI, magnetic resonance imaging; DWI, diffusion-weighted imaging; CTA, computed tomography angiography; MRA, magnetic resonance angiography; DSA, digital subtraction angiography; eTICI, expanded Thrombolysis in Cerebral Infarction.
